# Co-occurring infections in cancer patients treated with checkpoint inhibitors significantly increase the risk of immune related adverse events

**DOI:** 10.1101/2024.02.14.24302840

**Authors:** Tigran Makunts, Siranuysh Grabska, Hovakim Grabski, Ruben Abagyan

## Abstract

Therapeutic antibodies designed to target immune checkpoint proteins such as PD-1, PD-L1, and CTLA-4 have been applied in the treatment of various tumor types, including small and non-small cell lung cancers, melanoma, renal cell carcinoma, and others. These treatments combat cancers by reactivating CD8 cytotoxic T-cells. Nevertheless, this unique targeted mode of action was found to be associated with a broader range of immune-related adverse events, irAEs, affecting multiple physiological systems. Depending on their severity, these irAEs often necessitate the suspension or discontinuation of treatment and, in rare instances, may lead to fatal consequences. In this study we investigated over eighty thousand adverse event reports of irAEs in patients treated with PD-1, PD-L1, and CTLA-4 inhibitors. FDA Adverse Event Reporting System MedWatch submissions were used as the data source. These therapeutics included pembrolizumab, nivolumab, cemiplimab, avelumab, durvalumab, atezolizumab, and ipilimumab. The data analysis of these reports revealed a statistically significant association of immune related adverse events, including serious and life-threatening events in patients who experienced infectious disease during treatment. Additionally, the association trend was preserved across all the three classes of checkpoint inhibitors and each of the seven individual therapeutic agent cohorts.

## Introduction

Cancer immunotherapy has gained recognition due to the success of diverse targeted immune checkpoint inhibitors (ICIs) in treating a wide range of malignancies, including small and non-small cell lung cancers, melanoma, renal cell carcinoma, and others. Cancer cells possess an ability to evade the immune system by hindering the activation of T-cells(1-3). These treatments combat cancers by reactivating CD8 cytotoxic T-cells. The discovery of this mechanism has spurred the exploration of novel approaches to obstruct immune checkpoint breaks and rekindle the immune response(1). In 2011, the first approved immunotherapy antibody, ipilimumab (Yervoy)(4), blocking the cytotoxic T-lymphocyte–associated antigen 4 (CTLA-4)(4). Antibodies targeting the programmed cell death protein 1 (PD-1) receptors, namely pembrolizumab (Keytruda) and nivolumab (Opdivo), and cemiplimab (Libtayo) were approved later. PD-1 ligand (PD-L1) blocking antibodies, including atezolizumab (Tecentriq), durvalumab (Imfinzi), and avelumab (Bavencio) (1, 5) were approved as well.

Treatment with checkpoint inhibitors has been associated with significant immune-related adverse events (irAEs) (6). Inhibiting immune checkpoints can lead to the activation of auto-reactive T-cells and consequently lead to various irAEs impacting many organ systems, including pulmonary (e.g. pneumonitis, sarcoidosis), cardiac (e.g. myocarditis, pericarditis), renal (e.g. nephritis), gastrointestinal (e.g. colitis, hepatitis) etc (6).

In a prior study analyzing KEYNOTE clinical trials data for pembrolizumab(7-9), we observed a statistically and clinically significant association between co-occurring infections and irAEs(10). This finding warranted further investigation using postmarketing safety data for all approved ICIs. For a thorough evaluation of this association, United States Food and Drug Administration (FDA) Adverse Event Reporting System (FAERS) postmarketing surveillance data was used as the data source. Given the differences in the mechanisms of action of the ICIs, it was not clear if the trend observed pembrolizumab would remain for the other ICIs, considering their individual design differences and different targets.

## Main text

### Methods

#### FDA adverse event reporting system (FAERS/AERS) and MedWatch

FAERS, which includes its historical version AERS, is a database of post-marketing safety surveillance reports and operates under the auspices of the United States Food and Drug Administration (FDA). Reporting of adverse events (AEs) and associated outcomes to FAERS/AERS primarily occurs through the MedWatch (forms 3500 and 3500A) (11). It includes voluntary submissions from consumers, legal representatives, physicians, pharmacists, nurses and other healthcare professionals. Reports initially submitted to the sponsor/vendor/manufacturer, are required by regulations to be submitted to FAERS/AERS as well.

At the time of the analysis, FAERS/AERS data set consisted of over nineteen million adverse event reports spanning from the first quarter of 2004 (which includes reports from 1990’s) to the second quarter of 2023. These reports formed the basis for a retrospective analysis of immune checkpoint inhibitor AE cases.

For online access to FAERS/AERS datasets, please refer to the following link: https://fis.fda.gov/extensions/FPD-QDE-FAERS/FPD-QDE-FAERS.html

All methods and procedures for data analysis adhered to established guidelines and regulations. Given that the study exclusively utilized publicly accessible data, and the FDA datasets analyzed had undergone review, deidentification, and release, there was no need for additional approval from institutional or licensing committees.

#### Data preparation

FAERS/AERS consolidates reports from both the United States and other countries, each with its distinct demographic formats and medication brand/generic names. Prior to the initiation of data collection and analysis, online drug databases including (12)ChEMBL, ZINC(13), PubChem(14), DrugBank(15) and National Library of Medicine(16) were employed to establish a comprehensive dictionary of various drug generic and brand names, to facilitate their translation into unique generic names. Each of the quarterly report data sets from FAERS/AERS consists of seven files containing common report IDs. To enhance uniformity across the datasets, each quarterly report set was downloaded in dollar-separated text format (.txt) and reformatted into single reports and extended with generic names. To create a standardized data table, missing columns in the FAERS/AERS dataset were introduced without specific values(17-19). The resulting comprehensive table contained a total of 19,609,804 adverse event reports, nearly 0.4% of which were duplicate reports and were removed prior to the analysis.

#### Cohort choice and study outcomes

Prior to analysis, FAERS/AERS database was queried for all the reports of approved PD-1, PD-L1, and CTLA-4 ICIs (n=233,915). The reports where an ICI was the only reported drug used, further referred to as *monotherapy* reports, were selected (n=80,927) and further separated into individual classes: PD-1 inhibitors (pembrolizumab; n=22,580, nivolumab; n=38,218, cemiplimab; n=680), PD-L1 inhibitors (avelumab; n=912, durvalumab; n=4,398, atezolizumab; n=5,310), and CTLA-4 inhibitor (ipilimumab; n=8,829). Each individual ICI cohort was further split into cases with co-occurring infections and cases without co-occurring infections. For clarity, only reports with a single treatment by a cancer immunotherapy agent were retained to rule out patients with pre-existing infectious diseases. There were over twenty thousand unique AE terms listed in FAERS/AERS. The following FDA Medical Queries (FMQs) were used to generate a list of preferred term codes to define the *infection (primary)* and *no infection (control)* cohorts: bacterial infection, fungal infection, opportunistic infection, renal and urinary infection, and viral infection. A total of 2,545 infections and related FMQ preferred terms (PT) codes(20) were used in this study.

#### Study outcomes

The primary measured study outcome was the difference in reported irAE frequencies for both groups (*infection vs. no infection*). The irAE PT codes were based on any system organ class AE terms of inflammatory nature. The following reports for infection-related inflammatory conditions (e.g. viral hepatitis, infectious colitis) were excluded from the irAE list due to their unrelated etiology/root cause. However, the immune related AEs characteristic of enhances auto-immune reactions were retained. Reporting odds ratio (ROR) disproportionality analysis was performed using their relative irAE reported frequencies and evaluating the 95% confidence intervals (CI) of the ROR values (see details in <u>Statistical Analysis section</u>).

#### Statistical Analysis

##### Descriptive statistics

Frequencies for each AE PT code were calculated by the equation:

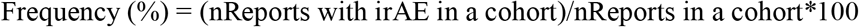

##### Comparative Statistics

AE report rates were compared via the Reporting Odds Ratio (ROR) analysis using the following equations:

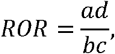

where:

*a* is the number of irAE cases in co-occurring infection group

*b* is the number of no irAE cases in co-occurring infection group

*c* is the number of irAE cases in control group

*d* is the number of no irAE cases in control group

Standard Error of Log Reporting Odds Ratio:

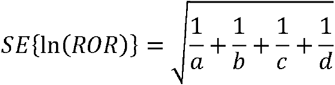

95% Confidence Interval:

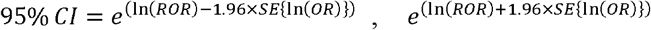

## Results

Monotherapy reports of seven studies immune checkpoint inhibitors contained a substantial number of reported irAEs: Pembrolizumab-23.3%, nivolumab-19.0%, cemiplimab-16.6%, avelumab-10.9%, durvalumab-29.7%, atezolizumab-14.3%, and ipilimumab-24.5%. Surprisingly, the irAE report occurrence in the ipilimumab cohort (CTLA-4 targeting antibody), did not differ significantly from PD-1 and PD-L1 cohorts (PD-1 total 20.6%, PD-L1 total 20.4% vs. CTLA-4 total 24.5%). Each individual *monotherapy* cohort was split into *co-occurring infection* and *no co-occuring infection* sub-cohorts, and the irAE reported frequencies were compared (Figure 1).

**Figure 1.**
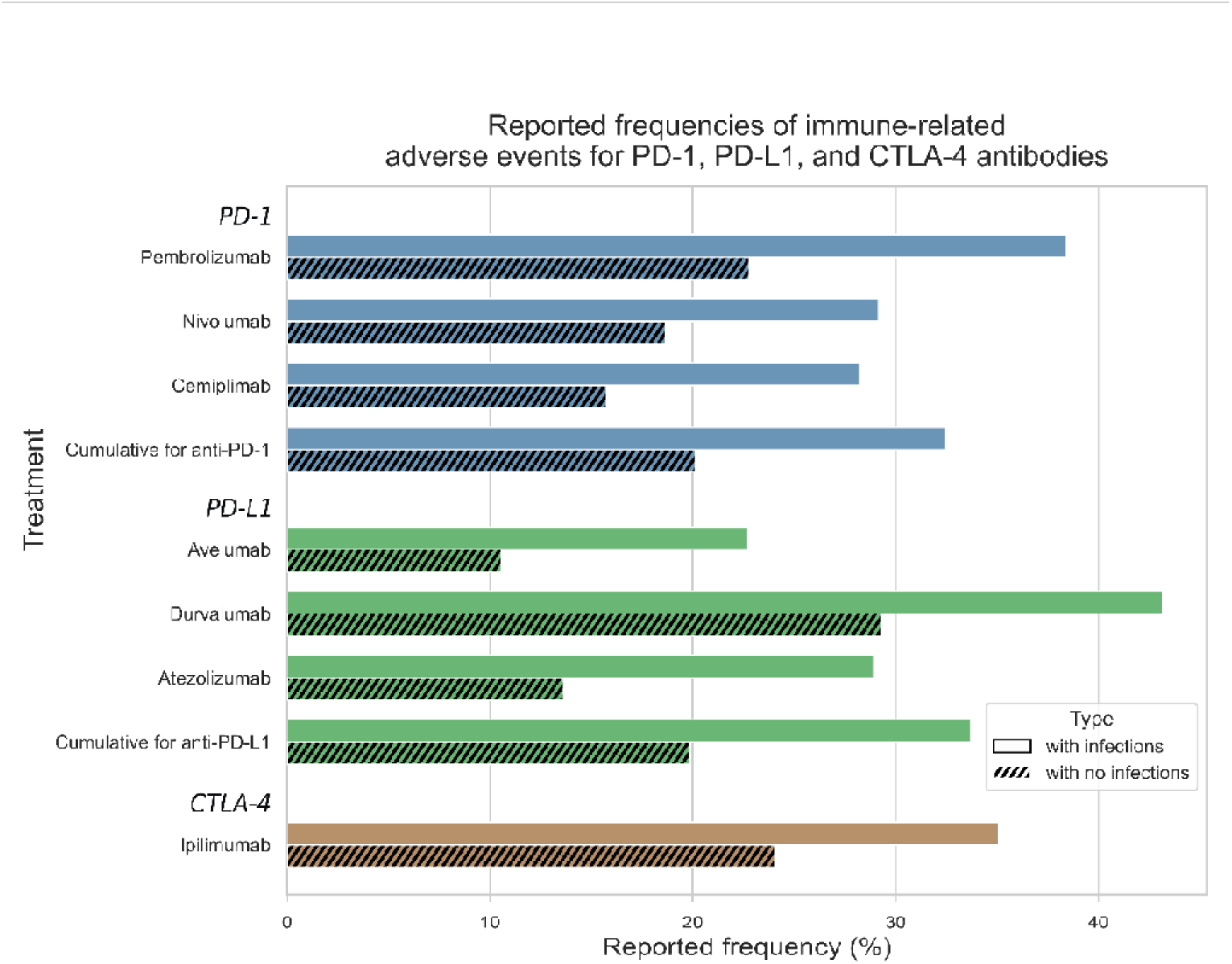
FAERS reported frequencies of irAE in patients administered ICIs with and without co-occurring infections.

The differences within each ICI class and individual ICI cohorts were *statistically significant* across all the ICI class cohorts. Patients administered ICIs who had a co-occurring infection had a higher risk of developing irAEs: pembrolizumab ROR 2.11 (95% CI [1.82, 2.45]), nivolumab 1.80 (1.59, 2.03), cemiplimab 2.10 (1.07, 4.14), durvalumab 1.83 (1.29, 2.60), atezolizumab 2.57 (1.89, 3.50), and ipilimumab 1.70 (1.33, 2.18). avelumab 2.49 (0.90, 6.91). For avelumab, although the trend was preserved, the risk was not statistically significant 2.49 (0.90, 6.91). Additionally, when analyzed cumulatively, each all the ICI classes had a statistically increased risk of ICIs when a *co-occurring infection* was present: PD-1 inhibitors 1.91 (1.74, 2.09), PD-L1 inhibitors 2.05 (1.64, 2.55), CTLA-4 inhibitor (ipilimumab) 1.70 (1.33, 2.18) (Figure 2)

**Figure 2.**
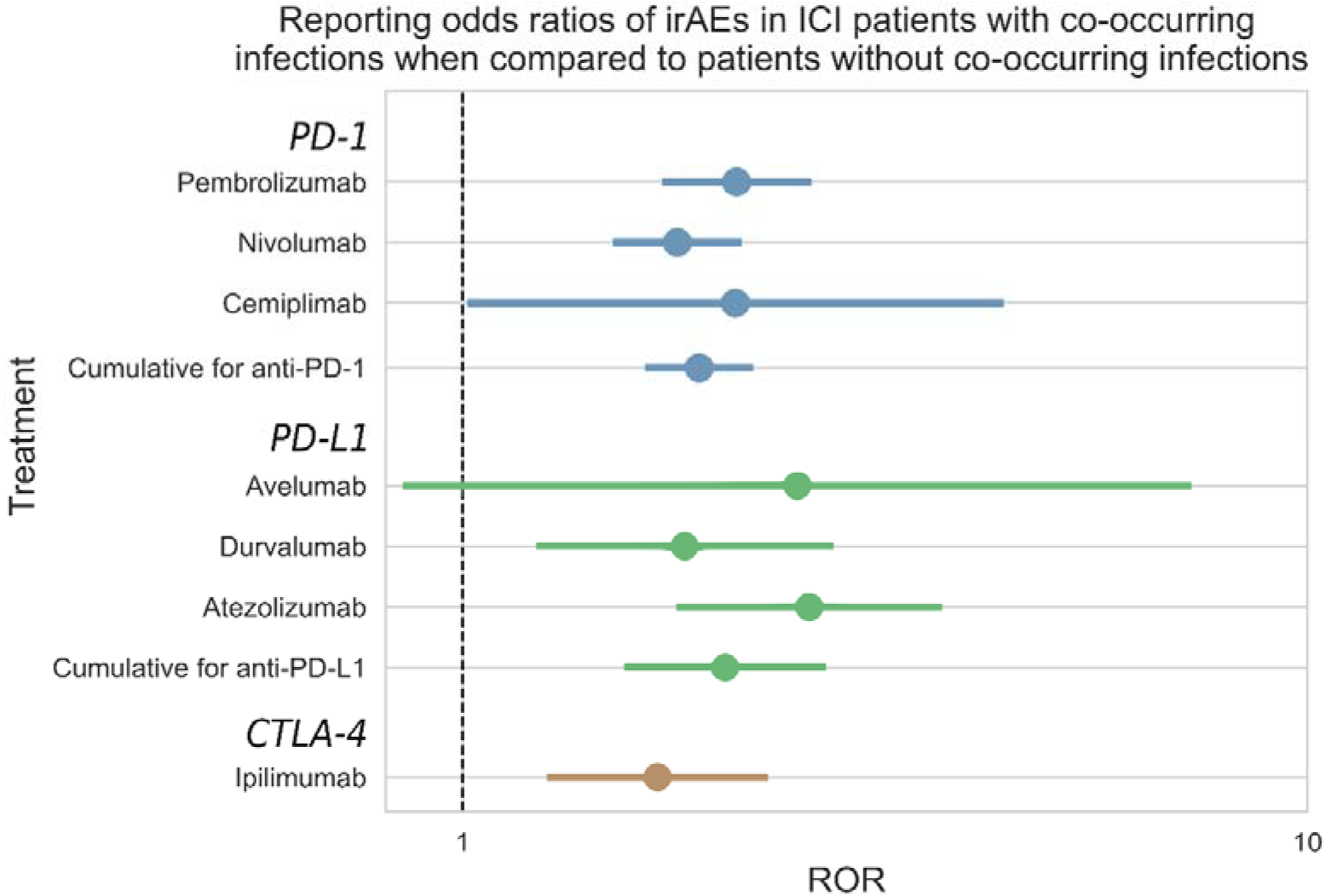
Reporting odds ratios (ROR) of irAEs in ICI patients with co-occurring infections when compared to patients without co-occurring infections. Ranges represent 95% confidence intervals (95% CI) (see Methods). A logarithmic X-axis shows odds ratios and their confidence intervals.

## Conclusion

In this study we analyzed over eighty thousand FAERS AE reports for individuals administered any of the seven approved ICIs as monotherapy for a broad range of malignancies for immune related adverse events. To the best of our knowledge, this is the first comprehensive examination of concurrent irAEs in subjects with and without co-occurring infections using population-scale postmarketing data.

The analysis revealed that risk of encountering an immune-related adverse event rose by 80-160% if cases where the individuals experienced a co-occurring infection during their treatment with immune checkpoint inhibitors. However, the relationship between the irAEs and the co-occurring infections was not straightforward, as there was no clear indication of causal relationship, due to the lack of any temporal details in the reports. This lack of longitudinal association made determining causality challenging. Nevertheless, the association was statistically significant and may have clinical implications for cancer patients at higher risk of immune related complications.

While there were a few case-reports and smaller-scale study publications discussing the connection between irAEs and infections(21, 22), with some authors attributing the related organ damage to irAE exacerbation during a concurrent infection(23), this association had not been previously quantified in large-scale studies(23). Although it seems intuitive that an infection could impact irAEs, the extent of this association had not previously been assessed. However, numerous studies have linked infections with autoimmune diseases (AD), which share similarities with irAEs in their manifestation, physiological profile, and molecular mechanisms involving innate and adaptive immune systems, including arthritis, autoimmune thyroiditis, colitis, and lupus(24-27) etc., where infection-related T-cell autoreactivity is the primary culprit. Mechanisms through which infectious agents might cause irAEs include cryptic antigen presentation, bystander activation, molecular mimicry, and epitope spreading(28).

Comparing the AD/infection with the ICI irAE/infection relationship made sense, considering the ICIs’ mechanism of action and the physiology behind ADs and irAEs.

In summary, we observed a statistically significant association between co-occurring infections and immune-related adverse events in cancer patients treated with PD-1 PD-L1, or CTLA-4 inhibitors and found the association to be statistically significant with 95% confidence intervals for the reporting odds ratios.

### Study Limitations

The causality between infections and irAEs was not clinically adjudicated due to the lack of comprehensive medical and laboratory records. However, the use of population scale postmarketing surveillance data provides a robust signal that may have clinical significance. Although we examined the data for concomitant medications and medical history to account for potential confounders, it is important to note that consumers and healthcare professionals often underreport over-the-counter medications, supplements, and even prescribed medications. Additionally, non-clinically significant medical events such as minor infections may go unreported, introducing uncertainties to the analysis due to the possible induction of autoimmunity by these agents(29, 30). IrAEs often go underreported due to the complexity of diagnosis, which may require an invasive procedure, often leading to mischaracterizations of these adverse events.

## Data Availability

The datasets analyzed for this study can be found in the FAERS database. For online access to FAERS/AERS datasets, please refer to the following link:
https://fis.fda.gov/extensions/FPD-QDE-FAERS/FPD-QDE-FAERS.html
The FDA Medical Queries tables were made available online in September 2022 and may be accessed at:
https://downloads.regulations.gov/FDA-2022-N-1961-0001/attachment_1.xlsm

https://fis.fda.gov/extensions/FPD-QDE-FAERS/FPD-QDE-FAERS.html

## Abbreviations

AE: adverse event
AERS: adverse event reporting system
AD: autoimmune disease
CI: confidence interval
CTLA-4: cytotoxic T-lymphocyte–associated antigen 4
FDA: Food and Drug Administration
FAERS: FDA Adverse Event Reporting System
FMQs: FDA medical queries
ICI: immune checkpoint inhibitors
irAE: immune-related adverse events
PD-1: programmed cell death protein 1
PD-L1: programmed cell death protein 1 ligand
ROR: Reporting odds ratios
PT: Preferred term

## Declarations

## Acknowledgments

We thank Dr. Da Shi for contributions to processing the FAERS/AERS data files and supporting the computer environment. We are also thankful to Dr. Isaac V. Cohen for the helpful discussions. We also thank the ADVANCE Research program of the Foundation for Armenian Science and Technology.

## Author contributions

SG and HG performed the experiments, RA and TM designed the study and, TM, SG, HG, and RA drafted the manuscript and reviewed the final version. RA processed the data set. All authors read and approved the submitted version.

## Conflicts of interest

The authors declare that they have no conflicts of financial or non-financial interest.

## Ethical approval

Not Applicable. The data sets available online to the public are de-identified. Institutional Review Board requirements do not apply under 45 CFR 6.102. There was no direct human participation in the study. All experiments were performed in accordance with relevant guidelines and regulations.

## Consent to participate

Not applicable

## Consent to publication

Not applicable

## Availability of data and materials

The datasets analyzed for this study can be found in the FAERS database. For online access to FAERS/AERS datasets, please refer to the following link: https://fis.fda.gov/extensions/FPD-QDE-FAERS/FPD-QDE-FAERS.html

The FDA Medical Queries tables were made available online in September 2022 and may be accessed at: https://downloads.regulations.gov/FDA-2022-N-1961-0001/attachment_1.xlsm

## Funding

The study was funded in part by NIH R35GM131881. The study was in part funded by FAST Armenia.

## Notes

### Competing Interest Statement

The authors have declared no competing interest.

